# Assessing germline mutational profile and its clinicopathological associations in Triple Negative Breast Cancer

**DOI:** 10.1101/2022.05.31.22275080

**Authors:** Jisha John, Ashwini Bapat, Siddharth Gahlaut, Naveen Luke, Rahul Kumar, Yashaswi Thakur, Aishwarya Konnur, Namrata Namewar, Ruhi Reddy, Shaheen Shaikh, Rituja Banale, Sanket Nagarkar, Smeeta Nare, George Thomas, Laleh Busheri, Asha Reddy, Devaki Kelkar, Santosh Dixit, Ashraf ul Mannan, Radhakrishnan Sabarinathan, Selvi Radhakrishna, Rupa Mishra, Chaitanyanand B Koppiker

**Author notes:** **Corresponding author** Chaitanyanand B. Koppiker M.D., Medical Director, Orchids Breast Health, Pune, India Managing Trustee, Prashanti Cancer Care Mission, Pune, India, Research Lead, Centre for Translational Cancer Research (CTCR), Pune India, Honorary Associate Professor, University of East Anglia, UK Director, Oncosciences, Jehangir Hospital, Pune, India Founding Director, BreastGlobal Network, Board of Directors and Visiting Faculty, School of Oncoplastic Surgery, CA, USA Member, Board of Directors, Indian Cancer Genome Atlas (ICGA), Contact. These Authors have contributed equally (Ashwini Bapat, Siddharth Gahlaut).

## Abstract

**Purpose:** Breast cancer is the most common cancer in Indian women with a high incidence of triple negative breast cancer (TNBC). The high TNBC prevalence (>25%) in India remains a challenge in clinical management. Association of germline BRCA1/2 mutations in TNBCs is well-established as a predisposing factor for hereditary breast cancer risk. These studies are however predominantly representative of western population. Therefore, we investigated germline profiles of multi-institutional cohort of TNBC patients in India

**Methods:** Using a multi-gene NGS (next-generation sequencing) panel of 26 ACMG recommended genes associated with inherited cancers.

**Results:** In our study cohort of 193 TNBC patients, we identified 57 pathogenic mutations of which *BRCA1* (71.93%) and *BRCA2* (14.03%) were most commonly mutated. Additionally, 8 pathogenic mutations were identified in non-BRCA genes associated with the HR pathway. 10 novel mutations were identified in 3 genes namely *BRCA1*, *BRCA2* and *PALB2*. Comparison of allele-frequency with the global databases like TCGA (The Cancer Genome Atlas), gnomAD and Genome Asia 100K indicated that the novel mutations were unique. Furthermore, we identified 48 variants of uncertain significance (VUS) (24.9%).

**Conclusions:** Our study confirms the major proportion of mutations in *BRCA1/2* genes in TNBCs in India. Interestingly, a higher proportion of VUS were found in the non-BRCA genes compared to BRCA1/2 emphasizing the need for functional studies of the non-BRCA genes. Additionally, large scale studies are also warranted to elucidate the landscape of germline mutations relevant to the Indian population and their probable clinical implications.

## Introduction

Breast cancer remains the most frequent malignancy in women globally with the highest incident rate[1]. It is a molecularly heterogeneous disease and increasing understanding of this heterogeneity has driven therapeutic management strategies over the past decade.

On the basis of molecular characteristics, breast cancers are classified into hormone receptor (estrogen receptor-ER and/or progesterone receptor-PR) positive tumors, human epidermal growth factor receptor 2 (HER2) positive, and triple negative breast cancers (TNBC) wherein the tumors do not express ER, PR and HER2. TNBCs account for 12-20% of all breast malignancies with most frequent occurrences in premenopausal women (age <40). Moreover, TNBCs have been observed to be more prevalent in African-American and South Asian women.

In India, TNBCs occur with a much higher prevalence (25-30%) compared to the western world (10-20%)[2], [3]. Since TNBCs lack targeted therapy and also exhibit a higher rate of recurrence, combined with delayed diagnosis and drug resistance, TNBCs remain a prominent challenge for breast cancer treatment in India. Interestingly, mutations in *BRCA1/2* genes associated with hereditary breast and ovarian cancers (HBOCs) are often observed at high frequency in TNBCs[4]. At about 10-20%, TNBCs harbor pathogenic *BRCA1* mutations with highest frequency out of all molecular subtypes[5], [6]. Corollary to this, is that about 60-80% of *BRCA1* mutation carriers have been shown to be at risk of developing TNBC over lifetime[7]–[9]. In addition to *BRCA1/2*, other genes involved in the DNA damage response pathway are also known to contribute to increased risk of hereditary breast and ovarian cancer (HBOC) syndrome. Founder mutations, mutations prevalent in individuals of a specific ancestry have been identified in *BRCA1/2*. International guidelines for genetic testing consider all these factors and recommend individuals with strong family history and TNBCs below 60 years to undergo genetic testing. Genetic screening programs are highly effective in identifying individuals with greater risk of developing cancer. Considering the correlation between *BRCA1/2* mutations and TNBCs, genetic screens can be potentially useful to distinguish individuals at high risk of developing TNBCs.

However few studies have elucidated the prevalence of mutations in the HBOC genes especially in the context of the Indian population. An understanding of the mutational profile of *BRCA1/2* and other HBOC genes and their clinical impact may have far reaching effects in mitigating life-threatening consequences of TNBCs in India. With this background we sought to assess the germline profiles of TNBC patients and its probable association with clinicopathological parameters. We demonstrate the utility of multi-gene panel testing to determine prevalence of BRCA and non-BRCA gene mutations and have also identified a few novel pathogenic mutations and several variants of uncertain significance (VUS) in our cohort.

### Methodology Clinical Management

Breast cancer diagnosis was based upon clinical examination and radiological evaluation of breast and axilla using Full Field Digital Mammography (FFDM) with 3-D Tomosynthesis (Siemens Mammomat Inspiration^TM^) and Ultrasonography (Siemens Acuson S2000^TM^). Histopathological studies on tru-cut biopsy samples (majority of cases) or vacuum assisted biopsy (for index tumors, Encor Ultra^TM^) samples were performed for confirming diagnosis of breast carcinoma. Similarly, ultrasonography and fine-needle aspiration cytology was used for investigating axillary lymph node metastasis. The breast cancer diagnosis was thus based on standard triple assessment using mammography/USG and core biopsy for HPE and IHC. Confirmed breast cancer cases underwent breast surgery at a network hospital site. The oncologic management with chemo-radiation protocols was undertaken by a multidisciplinary clinical team in accordance with the current NCCN guidelines.

### Eligibility

We conducted a retrospective cohort study on 193 TNBC patients with clinicopathological information, 133 from Prashanti Cancer Care Mission Pune and 60 from Chennai Breast Center over a period of January 2010 to January 2020. The triple negative status was confirmed by IHC for all 193 patients from a local laboratory. Personal medical history, family history and pedigree information was collected. Clinicopathological data including age at diagnosis, menopausal status, type and grade, axillary nodal status, NACT, surgery and post-surgery details, oncological follow up was collected from the clinic medical records. Germline testing was performed at a partner CAP accredited laboratory. Written informed consent was obtained from all patients. The study was approved by the Institutional Ethics Committee of PCCM (DCGI/CDSCO Registration Number: ECR/298/Indt/MH/2018. Ethics approval was also obtained from the Institutional Review Board of Chennai Breast Centre.

### Genetic testing - Next generation sequencing (NGS)

Genomic deoxyribonucleic acid (DNA) was extracted from blood/saliva sample using standardized methodology (PrepIT-L2P kit from DNA Genotek, Canada) or QIAamp DNA Mini Kit (Qiagen, Germany) or the Nucleospin kit (Macherey–Nagel, Germany) and quantified by Qubit fluorimeter (Life Technologies). The test included genes recommended by ACMG (the American College of Medical Genetics and Genomics) and associated with commonly inherited cancers (breast, colorectal, endocrine, gynecological, melanoma, nervous system, pancreatic and prostate) as well as several rare cancers. NGS covering the ACMG recommended genes was performed for the index patient. The sequencing libraries were prepared from extracted gDNA by fragmentation, tagmentation, adaptor ligation, amplification, enrichment and hybridization. The library was quantified to load on the NGS platform, Illumina MiSeq/NextSeq. The genetic variations were identified by using the STRAND® NGS software and interpreted using the StrandOmics™ platform. The reads from the FASTQ files were aligned against the whole genome build hg19 reference using STRAND® NGS analysis pipeline. Variant annotation and prioritization was done by automated pipelines in StrandOmics, a clinical genomics interpretation and reporting platform developed in-house.

The StrandOmics Variant Annotation engine includes algorithms to identify variant impact on gene using both public content (ClinVar, HPO, links to dbSNP, 1000 Genomes, Exome Variant Server, and five in-silico predictors - FATHMM, LRT, Mutation Assessor, Mutation Taster, and SIFT) and proprietary content (curated variant records).

In addition to the main genes, ‘pathogenic’ and ‘likely pathogenic’ variants are investigated for secondary findings in the genes present in the Strand Germline Cancer Test.

### Bioinformatics and Data Mining

The description of the sequence variants is in accordance with the guidelines of the HGVS (Human Genome Variation Society) nomenclature. Standardizing the interpretation and reporting of genomic results was done in concordance with the recommendation of the ACMG and the Association for Molecular Pathology (AMP).

Publicly accessible variant databases such as ClinVar, BRCA Exchange, dbSNP, BIC; Flossies (FL); Genome Nexus; Franklin (Genoox) were consulted for the clinical classification of the identified pathogenic variants. The variants were also checked against the list of variants from TCGA somatic and germline studies to identify if the variants identified in our study were previously reported or novel. The identified novel mutations were also checked in gnomAD and GenomeAsia 100K to determine if these have been previously reported. In silico analysis of the identified VUS was performed using predictors, such as A-GVGD, SIFT, Polyphen, Mutation Taster, Likelihood-ratio test, Mutation Assessor, FATHMM and Pathogenicity Calculator. Figure 1 is a schematic representation of the study design and workflow.

**Figure 1:**
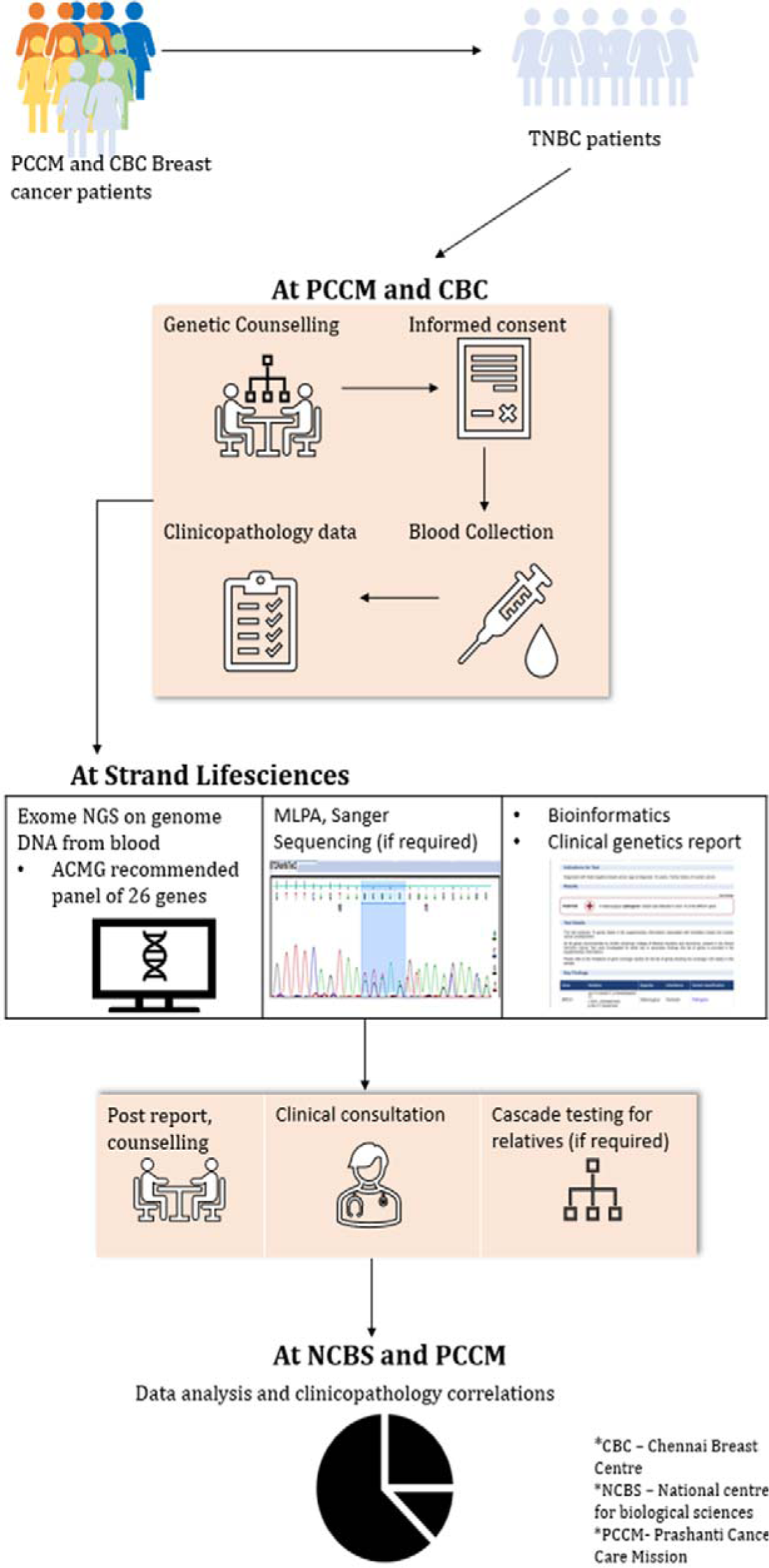
Schematic representation of patient recruitment and study design.

### Data analysis and Statistics: Chi-square statistical analysis

Differences between patient cohorts harboring pathogenic mutation, VUS mutation, and no mutation with regard to 17 clinicopathologic parameters were examined using Chi-square (χ2) test. p-value of <0.05 was considered statistically significant. The Python scipy stats library was used for statistical calculation.

## Results

The Clinicopathological parameters of the population included in the study are detailed in Table 1.

**Table 1:** Demographic and Clinicopathological parameters.

| Variables | Count (%) | total |
| --- | --- | --- |
| **Age at Diagnosis** |  |  |
| <40 | 60 (31.1%) | 193 |
| 40-50 | 60 (31.1%) |  |
| 50-60 | 40 (20.7 %) |  |
| >60 | 24 (12.4%) |  |
| NA | 9 (4.7%) |  |
| **Menopausal status at Diagnosis** |  |  |
| Pre-menopausal | 106 (54.9%) | 193 |
| Post-menopausal | 73 (37.8%) |  |
| Peri-menopausal | 2 (1.0%) |  |
| Post-menopausal (Hysterectomy) | 3 (1.6%) |  |
| NA | 9 (4.7%) |  |
| **Tumor Size** |  |  |
| ≤2 | 39 (20.2 %) | 193 |
| >2 | 131 (67.9 %) |  |
| NA | 23 (11.9%) |  |
| **Tumor Grade** |  |  |
| IDC I | 3 (1.6%) | 193 |
| IDC II | 42 (21.8%) |  |
| IDC III | 85 (44%) |  |
| IDC uncategorised | 13 (6.7%) |  |
| IDC + DCIS | 1 (0.5 %) |  |
| others (DCIS, ILC, Medullary carcinoma, papillary carcinoma) | 33 (17.1 %) |  |
| Bilateral | 4 (2.1 %) |  |
| NA | 12 (6.2 %) |  |
| **Recurrence** |  |  |
| Yes | 20 (10.4 %) | 193 |
| No | 160 (82.9 %) |  |
| NA | 13 (6.7) |  |
| **Metastasis** |  |  |
| Yes | 10 (5.2 %) | 193 |
| No | 101 (52.3 %) |  |
| NA | 82 (42.5 %) |  |
| **Survival Status** |  |  |
| Survivor | 173 (89.6 %) | 193 |
| Deceased | 4 (2.1 %) |  |
| Loss to follow up | 16 (8.3 %) |  |
| **Family History** |  |  |
| Sporadic | 97 (52.2 %) | 193 |
| Familial | 17 (8.8 %) |  |
| HBOC | 75 (38.9 %) |  |
| NA | 4 (2.1 %) |  |

### Mutational Spectrum

Germline mutation screening in genes recommended by ACMG identified 105 mutations in 94 patients. Among these 57/105 (54.28%) pathogenic mutations were detected across high, moderate and low penetrance ACMG recommended genes. Of these, the most frequently mutated genes were *BRCA1* (41/57, 71.93%) and *BRCA2* (8/57, 14.03%). The diagnostic yield of *BRCA1* was 21.24% (41/193), higher than other genes included in the panel, while that of BRCA2 was 4.14%.

Eight pathogenic mutations were detected in the non-BRCA predisposing genes (4.14%). The non-BRCA genes with mutations included *ATM*, *MLH1*, *MSH2*, PALB2, CHEK2 and *TP53* genes. Interestingly, 11 individuals were found to harbour mutations in two genes among which 3 individuals harboured pathogenic mutations in two cancer predisposing genes. Table 2 elaborates the distribution of the mutations in the cohort and Table 3 shows the mutational status of the 11 patients harbouring two mutations.

**Table 2:**
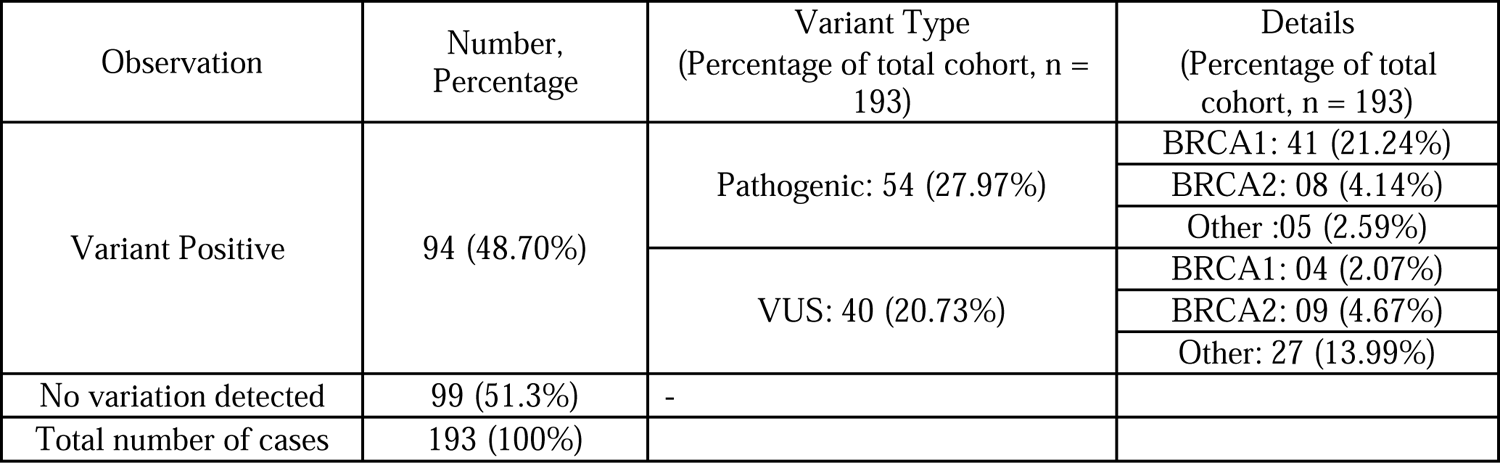
Distribution of mutations in the cohort.

| Observation | Number, Percentage | Variant Type<br>(Percentage of total cohort, n = 193) | Details<br>(Percentage of total cohort, n = 193) |
| --- | --- | --- | --- |
| Variant Positive | 94 (48.70%) | Pathogenic: 54 (27.97%) | BRCA1: 41 (21.24%) |
|  |  |  | BRCA2: 08 (4.14%) |
|  |  |  | Other :05 (2.59%) |
|  |  | VUS: 40 (20.73%) | BRCA1: 04 (2.07%) |
|  |  |  | BRCA2: 09 (4.67%) |
|  |  |  | Other: 27 (13.99%) |
| No variation detected | 99 (51.3%) | - |  |
| Total number of cases | 193 (100%) |  |  |

**Table 3:** Mutational status of 11 patients with more than one mutation in breast cancer predisposing genes.

| Strand ID | Gene | HGVS | Variant Classification |
| --- | --- | --- | --- |
| 1 | ATM | c.5631_5635delinsA | Pathogenic |
|  | MLH1 | c.306G>T | Pathogenic |
| 2 | ATM | c.8850+1G>A | Pathogenic |
|  | PALB2 | c.1447_1448delTC | Pathogenic |
| 3 | BRCA1 | c.2269delG | Pathogenic |
|  | MSH2 | c.1801C>T | Pathogenic |
| 4 | CHEK2 | c.733A>T | likely pathogenic |
|  | CDH1 | c.488G>C | VUS |
| 5 | ATM | c.3201C>G | VUS |
|  | NBN | c.251A>G | VUS |
| 6 | ATM | c.3899A>G | VUS |
|  | CHEK2 | c.937G>A | VUS |
| 7 | ATM | c.(8850+1_8851-1)_(*1_?)dup(duplication of exons 62-63) | VUS |
|  | NBN | c.485T>C | VUS |
| 8 | BRCA1 | c.4699G>A | VUS |
|  | NBN | c.547G>C | VUS |
| 9 | BRCA2 | c.1714G>A | VUS |
|  | PALB2 | c.3431T>C | VUS |
| 10 | BRCA2 | c.8471G>T | VUS |
|  | RAD51D | c.541G>A | VUS |
| 11 | MSH2 | c.1601G>A | VUS |
|  | STK11 | c.974G>C | VUS |

### Novel Mutations

10 novel mutations were detected in BRCA1, BRCA2 and PALB2 genes. 5 (45.45%) novel pathogenic variants detected in BRCA1 were exon 5 c.255_256delinsT, exon 7 and intron 7 c.536_547+166delinsT, exon 10-11 c.(670+1_671-1)_(4185+1_4186-1), exon 14 c.4658delT and exon 15 c.4831delG. Exon 15 c.4831delG was reported in 2 patients in the cohort.

BRCA2 exon 10 c.1761_1762insTA, exon 11 c.3663delT and exon 11 c.3730dupA were the 3 (27.27%) novel mutations found in the BRCA2 gene. 2 (18.18%) novel pathogenic mutations in PALB2 were found in exon 4 c.1447_1448delTC and exon 5 c.2398_2417dup.

### Recurrent Mutations

Exons 2, 10 and 11 in the BRCA1 gene and exon 11 in the BRCA2 gene were most frequently mutated regions. Of note, the c.68_69delAG, a founder Jewish Ashkenazi mutation, was detected in 12 patients (12/57, 21.05%) and was the most common pathogenic variant detected. Two pathogenic mutations were found in ATM and PALB2 respectively whereas one pathogenic mutation was detected in each of the MLH1, MSH2 and TP53 gene. The most common type of mutation in BRCA1 was frameshift (31/41) followed by splicogenic variants that were found in 6 patients. Exon 2 deletion, exon 10-11 deletion, exon 22-23 deletion and only 1 nonsense mutation was reported within the BRCA1 mutations. Similarly, frameshift mutations (6/8), 1/8 nonsense mutation and 1/8 duplication mutation were detected in BRCA2. The spectrum of mutations and their distributions in BRCA1 and BRCA2 is shown in Figure 2. Supplementary Figure 1 shows the position of these mutations on the BRCA1 and BRCA2 protein sequences.

**Figure 2:**
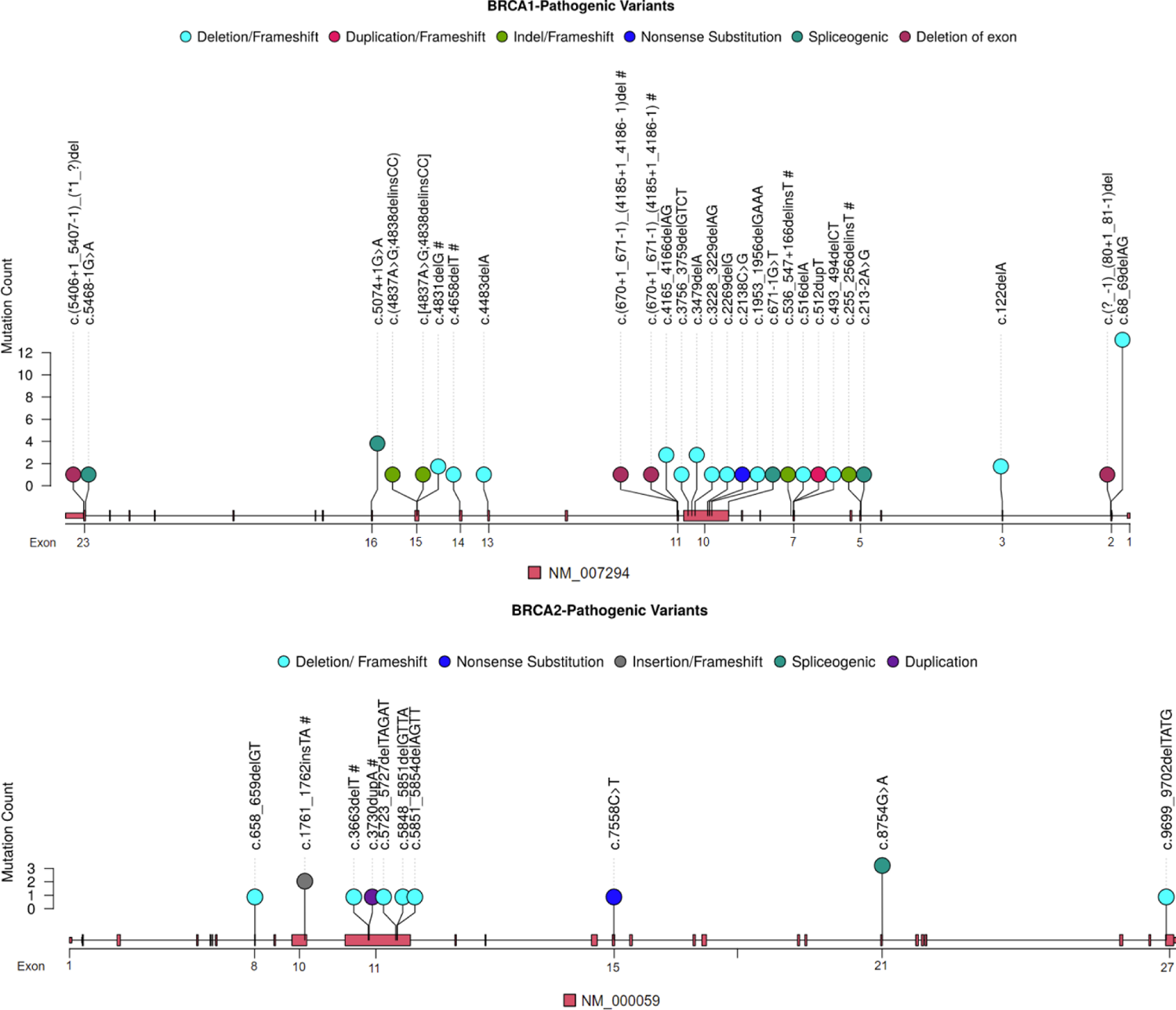
Distribution of the mutational spectrum in BRCA1 and BRCA2 genes. The lollipop plot represents the positions of the different pathogenic variants that were observed in both intron and exon regions of the BRCA1 an BRCA2 transcripts. The height of the lollipop indicates the number of observed mutations in the indicated positio (includes cascade testing). The color of the lollipop indicates the type of mutation observed. The shaded rectangles in the transcript indicate the different exons and the numerical labels indicate the corresponding exon number. The novel mutations identified in this study in these two transcripts are indicated by a #.

### Variants of uncertain significance (VUS)

A large proportion of VUS were detected in non-BRCA genes. Figure 3 illustrates the distribution of pathogenic and VUS mutations in BRCA1/2 and non-BRCA genes. 48/105 (45.72%) VUS were detected during the screening of 193 patients. Among these 13/48 (28.88%) VUS were found in BRCA1/2 gene (8.33% in BRCA1 and 18.75% in BRCA2). 72.92% VUS detected were in non-BRCA genes (35/48) included APC, ATM, BRIP1, CDH1, CHEK2, MLH1, MSH2, MSH6, NBN, NF1, PALB2, PMS2, PTEN, RAD51D, STK11, TP53, VHL genes. There were 4 missense mutations identified in ATM, 3 in MSH6, 2 in PMS2 and 1 each in the PALB2, TP53, MLH1, MSH2 genes.

**Figure 3:**
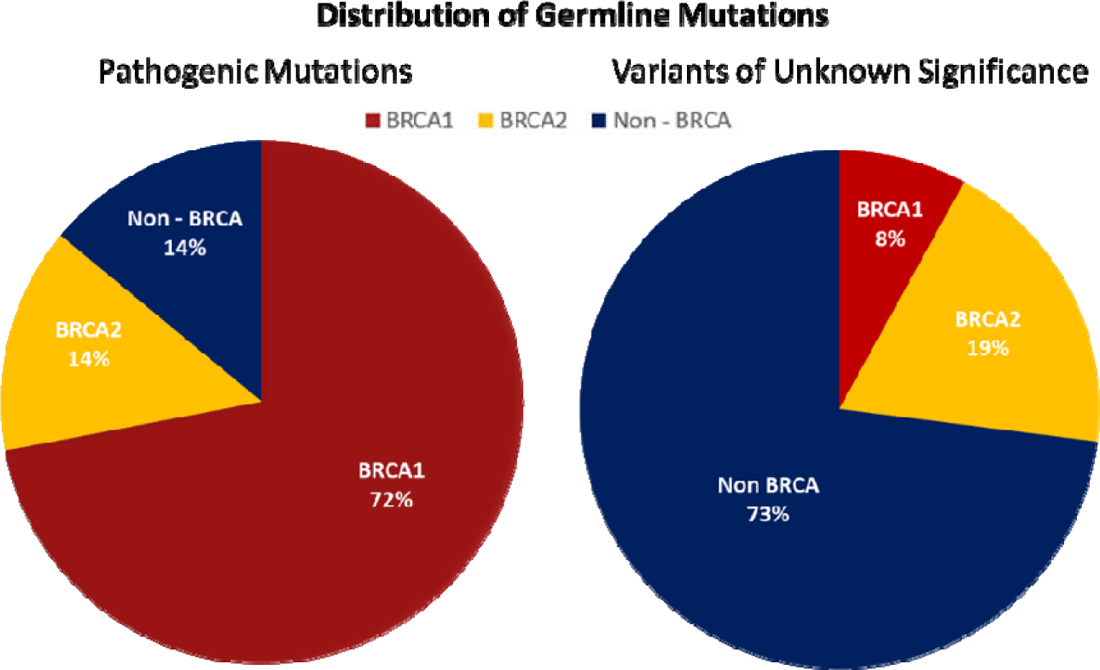
The figure depicts the distribution of germline pathogenic mutations and variants of unknown significance in BRCA1, BRCA2 and the non-BRCA genes

Family history is known to be a significant contributor to breast cancer risk. Family history wa determined based on the pedigree of the individual patients[7], [10]–[12]. Figure 4 depicts the distribution of germline mutations based on family history as determined by pedigree analysis. Among the 99 patients who tested negative for mutations, 29 reported Family History for HBOC while 9 had family history for cancer. Of the 4 patients harbouring *BRCA1* VUS 2 patients reported a HBOC family history whereas 2 had no cancer history in the family. In BRCA2 VUS patients, 4 of the 9 reported family history, 3 of which had HBOC family history. Interestingly, among patients with non-BRCA VUS mutations, 10 of the 27 reported HBOC family history while 1 patient had familial cancer history.

**Figure 4:**
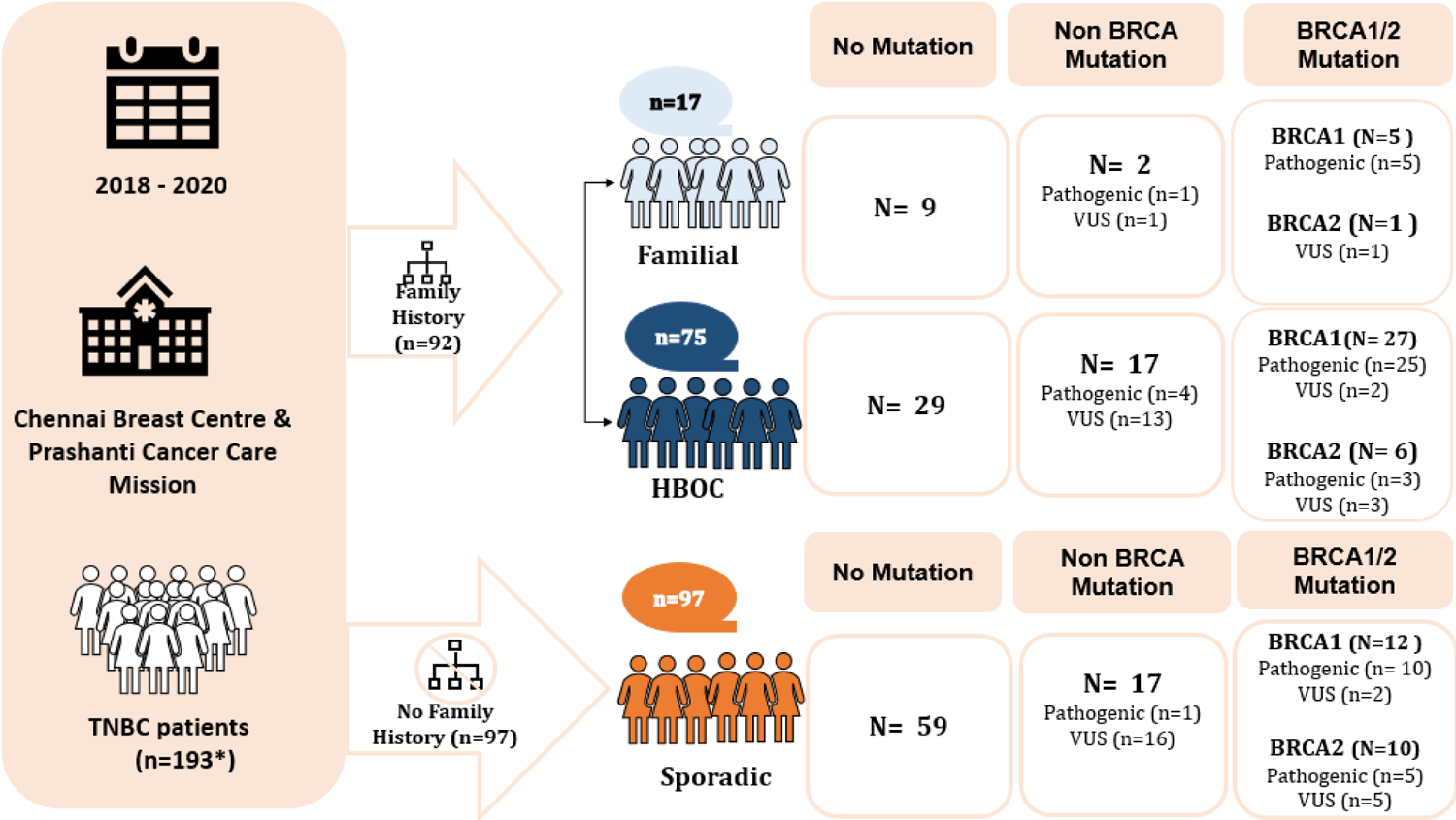
Distribution of Germline Mutations based on Family History: The graphic illustrates the distribution of family history in the study cohort. Among 193 patients, family history was unavailable for 4 patients. Additionally, 4 individuals with a family history of HBOC harbored two mutations (BRCA1 pathogenic + non-BRCA (MSH2) pathogenic (n=1); BRCA2 VUS + Non-BRCA (PALB2, RAD51D) VUS (n=2); Non-BRCA (CHK2) pathogenic + Non-BRCA (CDH1) VUS (n=1) and 1 individual with no family history was found to harbor a BRCA1 VUS + Non-BRCA (NBN) VUS

### Clinicopathological Parameters and their correlation with germline mutational status

The median age at diagnosis for our cohort was 46 years. A significant difference in age at diagnosis was observed for BRCA pathogenic mutations with median age at diagnosis for BRCA pathogenic mutation carriers of 42 years A significant correlation was observed between presence of pathogenic mutations and family history a well as metastasis (Table 4). Interestingly, of 9 patients with age at diagnosis above 60 and no family history, 3 patients were identified harboring germline pathogenic mutations. Future studies with a larger cohort may help determine other parameters that may be associated with germline mutational status of HBOC predisposing genes.

**Table 4:** Chi-square correlations of germline mutational status and clinicopathological features. A significant correlation was observed between pathogenic germline mutations and metastasis and family history.

|  | Total Pathogenic (n=55) | Total VUS (n=44) | No mutation (n=99) | Total Pathogenic vs No mutations |
| --- | --- | --- | --- | --- |
| **Metastasis** |  |  |  | **0.000** |
| Yes | 2 | 5 | 5 |  |
| No | 31 | 7 | 1 |  |
| NA | 15 | 6 | 1 |  |
| **Survival status** |  |  |  | **0.974** |
| Survivor | 49 | 39 | 89 |  |
| Deceased | 1 | 1 | 2 |  |
| Loss to follow up | 5 | 4 | 8 |  |
| **Family history** |  |  |  | **0.001** |
| Sporadic | 16 | 23 | 59 |  |
| Familial | 6 | 2 | 9 |  |
| HBOC | 32 | 18 | 29 |  |
| NA | 1 | 1 | 2 |  |

## Discussion

Triple negative breast cancers are among the most aggressive subtypes of breast cancer, commonly associated with poor prognosis and poor survival. Germline mutations in breast cancer predisposing genes have been identified in TNBC patients. As per international guidelines, all TNBC patients below age 60 are advised to undergo genetic testing to determine presence of a hereditary cause of the disease. Large scale genetic studies conducted in western population have identified the presence of specific mutations in HBOC genes that contribute to increased risk of the disease. In addition to identifying high penetrance and low penetrance genes, these studies also demonstrate the frequent occurrence of specific mutations in populations with common ancestry. These founder mutations have been identified in BRCA genes for Icelandic, Ashkenazi Jews and French-Canadian populations[13], [14]. However, no such founder mutations have been identified for the Indian population. In the current study we have assessed the profile of germline mutations in TNBC patients from our cohort and discerned association of clinicopathological parameters with mutational status, if any.

Among the 193 triple negative breast cancer patients, 94 (48.70%) patients were found to harbour at-least one mutation in the breast cancer predisposing genes identified by multi-gene panel testing. Among the pathogenic mutations, BRCA1/2 mutations formed a major share. Interestingly, the cumulative diagnostic yield of the BRCA1/2 genes identified in the current study is comparable to a previous study published by Yadav et. al. in 2018, where 27% of the TNBC patients from a cohort of 266 harbored a pathogenic BRCA mutation[6]. A similar study by Singh et. al. in over 1000 unrelated breast/ovarian cancer patients identified similar trends with 84.9% of mutations identified in BRCA1/2 genes. Internationally, although a report from Pakistan shows similar trends, studies from other countries such as USA, Germany, Spain and South Africa show comparatively lower prevalence of BRCA1/2 mutations, with only 10-15% of mutations attributed to BRCA1/2 genes [5], [8], [15], [16]

The most recurrent mutation was the c.68_69delAG in BRCA1, the founder mutation in the Ashkenazi Jewish population, with 12 patients from our cohort harbouring this mutation. This mutation has been reported among HBOC patients across different ethnicities with varying frequencies. The mutation is predicted to cause truncation at the beginning of the zinc-binding region of the RING domain in BRCA1 and may contribute to cancer predisposition by inactivating the ubiquitin protein ligase activity[17]. Population studies carried out across the world have revealed that the c.68_69delAG mutation is likely around 2000 years old and predates the separation of Sephardi and Ashkenazi Jewish populations[18]. The presence of population mixing, or large-scale migration could be accountable for the high prevalence of this founder mutation within our cohort. In India, studies conducted so far report the occurrence of this mutation with frequency of 0.5%–4.1% in breast cancer cases in different states[19]–[23]. Further studies involving larger patient numbers are warranted in order to come up with a mutation hotspot panel specific to the Indian population as a first-tier testing.

The c.5074+1G>A in BRCA1 and c5631_5635delInsA in ATM were the second most frequent with 4 patients from the cohort harboring these mutations. The c.5074+1G>A in BRCA1 is a spliceogenic variant (reported in BIC as 5193G>A), found within intron 16 of BRCA1. It has been previously reported as a founder mutation in the Icelandic population, though the prevalence is extremely rare and only accounts for 1% of the breast/ovarian cancers in Iceland[24], [25]. This variant has been classified as pathogenic based on multifactorial likelihood models and functional assays, and is predicted to cause aberrant splicing, resulting in a loss-of-function protein that is subject to nonsense-mediated mRNA decay [26], [27]. Within the Indian population, this variant has been reported to occur at a low frequency in breast cancer patients[22], [28], [29]. Truncated protein products, caused as a result of frameshift mutations, constitute for ∼90% of the clinically significant entries reported in the Breast Cancer Information Core (BIC) database. Among BC patients, Exon 11 is reported to be the most predominant location for mutations (both the BRCA1 and BRCA2 genes) and accounts for nearly 50% of these BIC reported mutations[30]. A study by Bayraktar et. al. also reported Exon 11 to be the preferred location for both the BRCA1 (103/445, 23.2%), and BRCA2 (92/445, 20.7%) mutations[31].

The c5631_5635delInsA mutation in ATM is also a pathogenic frameshift mutation. However, large scale cohort studies on the prevalence of this mutation in the population are lacking. Mutations in the genes involved in the homologous recombination pathway have been implied to be one of the drivers of TNBCs[8]. The mutations in ATM, BARD1, BRIP1, PALB2, RAD50, RAD51C, RAD51D involved in homologous recombination have also been shown to confer 10% increased risk of developing breast and ovarian cancer[16]. In our study we observed 14% of the cases harboring pathogenic mutations in non-BRCA genes, similar to the report by Singh et. al. 2018[32]. However, Buys et.al. report 51% mutations in non-BRCA genes using multi-gene panel testing, implying a possible ethnic difference in the prevalence of mutations attributable to BRCA vs non-BRCA genes[33].

According to the latest NCCN guidelines, clinical management, screening protocols and risk mitigation strategies have been included for a few non-BRCA genes. Annual screening by mammogram with tomosynthesis or MRI with contrast starting at age 40 for CHEK2 carriers has been recommended by NCCN. Similarly, for CDH1 and NF1 carriers, NCCN has advised annual mammograms with tomosynthesis and breast MRI with contrast starting at age 30. For individuals harbouring a PALB2 mutation, increased surveillance, especially after 30 years in case of strong family history and risk reduction mastectomy has been recommended as per the latest NCCN guidelines[34]. Risk reduction salpingo-oophorectomy may also be discussed for BRIP1, RAD51C and RAD51D carriers as per the guidelines. However, unfortunately, no effect on the mortality rate has as yet been reported as a result of the aggressive surveillance protocols[35].

Interestingly, we identified 10 novel mutations across three genes which have not been previously reported in any of the databases. These novel mutations might be specific to the Indian population; 5 in BRCA1, 3 in BRCA2 and 2 in PALB2. Nine of the 10 novel mutations identified within our cohort are frameshift mutations, resulting in consequent premature termination of the protein and loss-of-function. The one remaining novel variant, exon 7 and intron 7 c.536_547+166delinsT, causes deletion of exon 10-11 of the BRCA1 gene, which might result in loss-of-function.

Inadequate information about VUS poses a challenge for counselling and adds complexity to the overall clinical management of patients[22]. The advent of NGS-based multi-gene panel testing has resulted in an increasing number of VUS being identified. The prevalence of these mutations in each cohort depends on the sequencing parameters since clear variance has been observed across different studies globally in terms of the frequency of VUS, ranging from 5-6% in US patients with European ancestry and about 21% among African American patients[32], [36]–[39]. The increasing number of VUS by NGS has also led to varied interpretations and counseling by clinicians and counselors. In the current study, while BRCA1/2 mutations accounted for the major share of the pathogenic mutations identified (86%), 73% of the VUS identified were attributable to non-BRCA genes, although BRCA2 accounted for the highest number of VUS (9 variants). Among the 48 VUS identified, the majority were missense variants that are considered clinically ambiguous due to their unclear biological significance and lack of interpretation. This further highlights the need for reclassification which will help ascertain their biological role if any through various functional and co-segregation studies with other pathogenic variants.

Identification of deleterious mutations in the cancer predisposing genes may alter the clinical management and help implement aggressive risk reduction strategies for recurrence[13], [33]. The identification of pathogenic variants in multiple genes might also aid in the formulation of future guidelines specific to the Indian population. The high prevalence of BRCA mutations in our cohort further highlights the importance of risk prediction testing (cascade testing). Identification of high-risk individuals that can be benefited from risk-reducing surgical options like risk-reducing mastectomy, risk-reducing salpingo-oophorectomy or aggressive screening programs for early diagnosis may help mitigate the burden of breast cancer in the country.

We also analyzed possible clinicopathological correlations with germline mutational status. Despite the smaller cohort, we observed significant correlation of family history and metastasis with pathogenic germline mutations compared to individuals harboring no mutations. Interestingly we also observed a lower median age at diagnosis for individuals harbouring germline pathogenic mutations in BRCA genes compared to the rest of the cohort. Large scale studies coupled with longer follow-up may help determine the specific clinical contribution of the germline mutations in breast cancer etiology.

## Supplementary Data

**Supplementary Figure 1.**
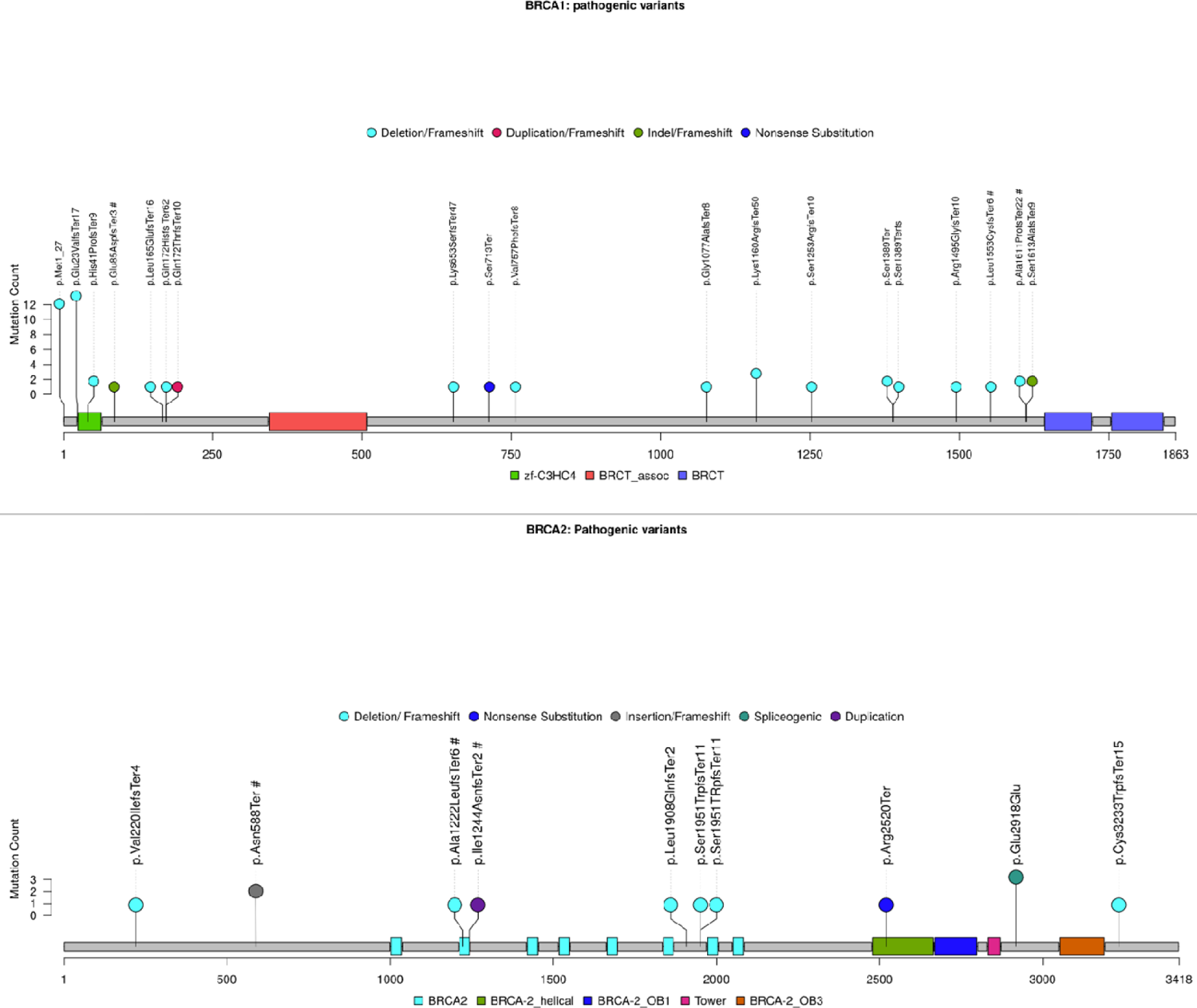
Lollipop plot representing the impact of mutations on the BRCA1 and BRCA2 protein sequence. The shaded boxes represent the different PFAM domains in the protein. The height of the lollipop indicates the number of times the indicated pathogenic variants were observed in this study while the color represents the type of mutation. The mutations identified exclusively in this study are indicated by a hash (#).

**Supplementary Figure 2.**
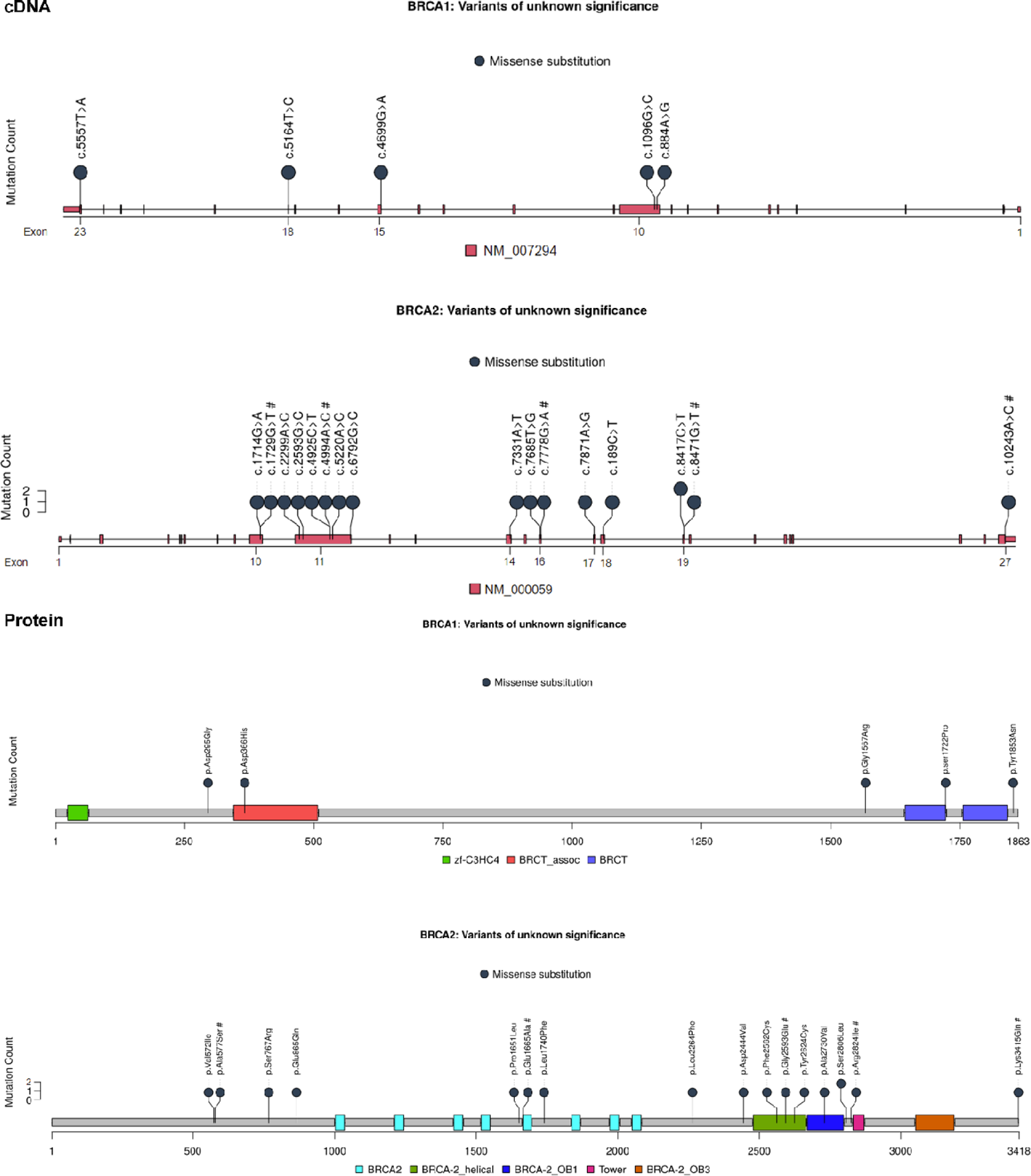
Lollipop plot representing the variants of unknown significance (VUS) in the transcript (upper panel) and its impact on the protein sequence (lower panel) for both BRCA1 and BRCA2 genes. The height of the lollipop indicates the number of times the indicated pathogenic variant were observed in this study while the color represents the type of mutation. The shaded boxes in the transcript represent exons while those in the protein sequence represent different PFAM domains. The mutations identified exclusively in this study are indicated by a hash (#).

## Data Availability

All data produced in the present work are contained in the manuscript.

## ACKNOWLEDGEMENT

The study authors would like to thank all participants who consented to participate in this study. We also acknowledge Bajaj Auto Ltd. for providing support to research activities at PCCM (Grant #PCCM0529 and Grant#PCCM GC2528). We are thankful to Dr Sneha Joshi, Dr. Madhura Kulkarni and Mr. Ankush Dewle for their support in preparation of this manuscript. The authors would also like to thank National Comprehensive Cancer Network (NCCN), Genetic/Familial High-Risk Assessment: Breast, Ovarian, and Pancreatic, Version 2.2021, NCCN (www.nccn.org) for the reference guidelines for genetic/familial breast cancer

